# Closing the Lung Cancer Screening Gap in FQHCs with AI-Powered Clinical Decision Support

**DOI:** 10.1101/2025.08.18.25332990

**Authors:** Arvil Dey

## Abstract

**Background:** Lung cancer remains the leading cause of cancer-related mortality in the United States, with screening adherence rates below 16% nationally and even lower among underserved populations [1–4]. Federally Qualified Health Centers (FQHCs), which serve over 30 million patients annually, face significant challenges in identifying screening-eligible individuals due to workforce constraints, manual workflows, and limited clinical decision support (CDS) infrastructure [5–7].

**Objective:** We developed and evaluated an AI-driven, natural language (NLP)-enhanced CDS platform designed to automate lung cancer screening eligibility determination and integrate seamlessly into the workflows of FQHCs.

**Methods:** The platform combines a deterministic rules engine, encoding the U.S. Preventive Services Task Force (USPSTF) 2021 screening criteria, with an NLP pipeline built using SciSpacy and MedSpaCy to extract smoking history details from unstructured clinical notes. Structured and unstructured data were unified using FHIR-compliant APIs to generate actionable screening flags within simulated FQHC workflows.

**Results:** In a synthetic cohort modeled on FQHC populations, the platform achieved precision, recall, and F1-scores above 0.90 for eligibility determination and reduced manual chart review workload by 62%. It generated clear, auditable recommendations aligned with Centers for Medicare & Medicaid Services (CMS) coverage policies and Healthcare Effectiveness Data and Information Set (HEDIS) quality measures, while identifying additional high-risk patients who may have been missed in manual reviews.

**Conclusion:** This work demonstrates the feasibility of an AI-powered CDS framework for improving lung cancer screening adherence in resource-limited primary care settings. Future development will focus on prospective validation, expansion to other preventive screenings and population health use cases, and integration of explainable AI to enhance clinical trust and scalability.

## INTRODUCTION

Lung cancer is the leading cause of cancer-related mortality in the United States, accounting for more deaths annually than breast, prostate, and colorectal cancers combined [1]. Nearly 236,000 new cases and approximately 130,000 deaths occur each year [1, 2]. Despite clear evidence supporting low-dose computed tomography (LDCT) for early detection, fewer than 16% of eligible Americans undergo screening [3], with particularly low participation among underserved populations [4].

Federally Qualified Health Centers (FQHCs), which provide care to more than 30 million predominantly low-income and rural patients [5], face systemic barriers such as workflow shortages, limited infrastructure, and manual, error-prone eligibility workflows. These constraints delay diagnoses, increase costs, and worsen outcomes, reinforcing disparities in cancer care [6, 7].

Existing clinical decision support (CDS) tools have not bridged this gap. Alert-heavy EHR systems often exacerbate clinician fatigue without sustainably improving guideline adherence [8], while AI-driven solutions typically require technical expertise and infrastructure that are impractical for safety-net providers [9].

To address these challenges, we developed Onqi Screening, an AI-enhanced CDS platform that automates lung cancer screening eligibility within FQHC workflows in alignment with the U.S. Preventive Services Task Force (USPSTF) and Centers for Medicare & Medicaid Services (CMS) guidelines [10, 11]. The platform is designed to reduce manual burden, improve equity in early cancer detection, and strengthen value-based care delivery [17].

This work not only demonstrates the feasibility of a scalable, real-world AI solution for resource-limited healthcare settings but also establishes a framework for extending the platform to other preventive screenings and population health interventions. In doing so, it aligns with national priorities such as the Cancer Moonshot [14] and Healthy People 2030 [15], positioning this approach as a critical step toward technology-enabled public health infrastructure capable of reducing disparities and improving outcomes at scale.

## METHODS

### Study Design and Overview

We conducted a retrospective, simulation-based study to evaluate an artificial intelligence (AI)-driven CDS prototype aimed at closing the lung cancer screening gap in FQHCs. The primary objective was to measure the prototype’s accuracy in automating screening eligibility determination. Secondary objectives included assessing its fit within existing clinical workflows and estimating its potential to reduce manual workload, improve screening adherence, and strengthen quality reporting.

The system was developed in alignment with the USPSTF guidelines [10] and the CMS National Coverage Determination [11]. We used an early version of the AI-driven screening platform developed by the authors (Onqi Screening), designed specifically to address operational barriers to screening in resource-constrained FQHC settings.

### Data Sources

The CDS prototype was developed and tested entirely on synthetic datasets. No patient records or restricted datasets were accessed in this study.

To ensure reproducibility, we generated structured and unstructured synthetic data as follows:

- **Structured data** (e.g., age, gender, smoking status, pack-years, quit-years) were generated programmatically using the Faker library (v24.9.0) in Python. Distributions were informed by publicly available epidemiologic statistics (for example, CDC [1], HRSA/FQHC population reports [5], USPSTF lung cancer screening guidelines [10]), so that the simulated population reflected higher tobacco use prevalence and socioeconomic risk factors typical of FQHCs.
- **Unstructured data** (clinical notes) were created synthetically using text templates modeled on documentation patterns from publicly available datasets such as MIMIC-III discharge summaries [20] and the i2b2 smoking challenge notes [21]. No original sentences were copied; instead, patterns and phrasing styles were adapted to simulate realistic provider documentation.

All generated data are reproducible using the scripts available in our public repository: https://github.com/adey1998/onqi-research.

This approach ensured that the CDS prototype could be evaluated in a controlled, reproducible environment while avoiding the use of identifiable patient records.

### Eligibility Criteria

Eligibility was determined according to USPSTF 2021 guidelines:

- Age 50-80 years
- ≥ 20 pack-years smoking history
- Current smoker, or former smoker who quit < 15 years ago

Patients outside these criteria were flagged as ineligible, with the reason for ineligibility recorded (e.g., insufficient pack-years, age outside range, quit ≥ 15 years ago).

### System Architecture

The CDS prototype was implemented using a modular, cloud-ready architecture designed for deployment on Google Cloud Platform (GCP). Although this evaluation was conducted in a local simulation environment, the architecture was engineered for scale, with planned deployment leveraging GCP services such as Cloud Run, Cloud SQL, and Cloud Monitoring.

The architecture comprised three primary layers:

1. **Data ingestion layer:** Retrieved structured and unstructured patient data through secure FHIR-compliant APIs, ensuring HIPAA-compliant handling of protected health information [18].
2. **Processing layer:** Applied a deterministic rules engine that encoded USPSTF lung cancer screening criteria to determine eligibility based on age, smoking history (pack-years), and quit-years. For unstructured data, the system incorporated a natural language processing (NLP) pipeline built with SciSpacy [12] and MedSpaCy [19]. This pipeline extracted smoking-related details (smoking status, pack-years, quit-years) from clinical notes, followed by regular expression-based normalization to ensure accuracy.
3. **Output layer:** Combined structured and extracted data to generate clear eligibility flags and actionable screening recommendations. These outputs were designed to integrate directly into clinical workflows using FHIR ServiceRequest resources, enabling seamless access for providers without increasing manual workload.

**Figure 1.**
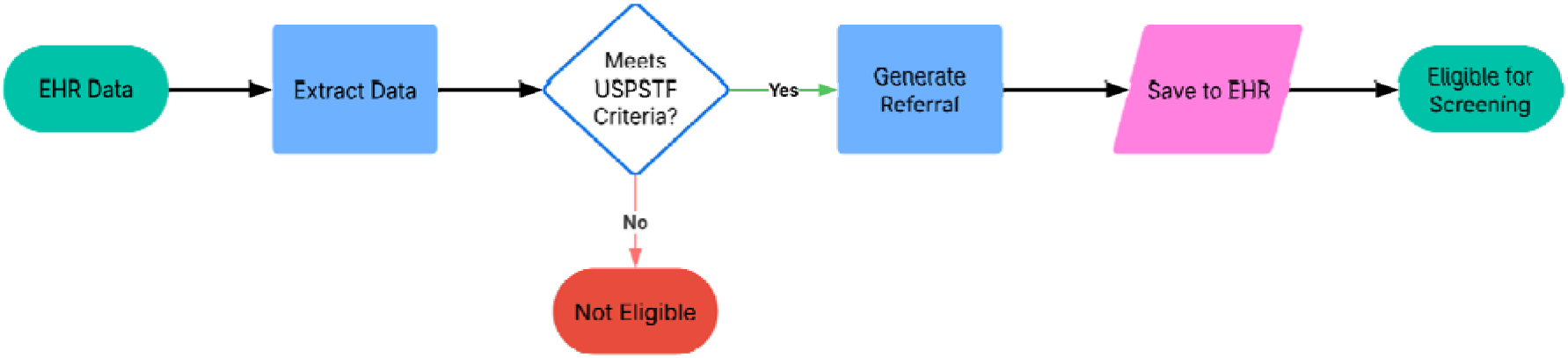
Eligibility engine logic for lung cancer screening. Deterministic rule-based pipeline applying USPSTF 2021 guidelines using age, pack-years, and quit-years derived from structured EHR data and NLP output.

**Figure 2.**
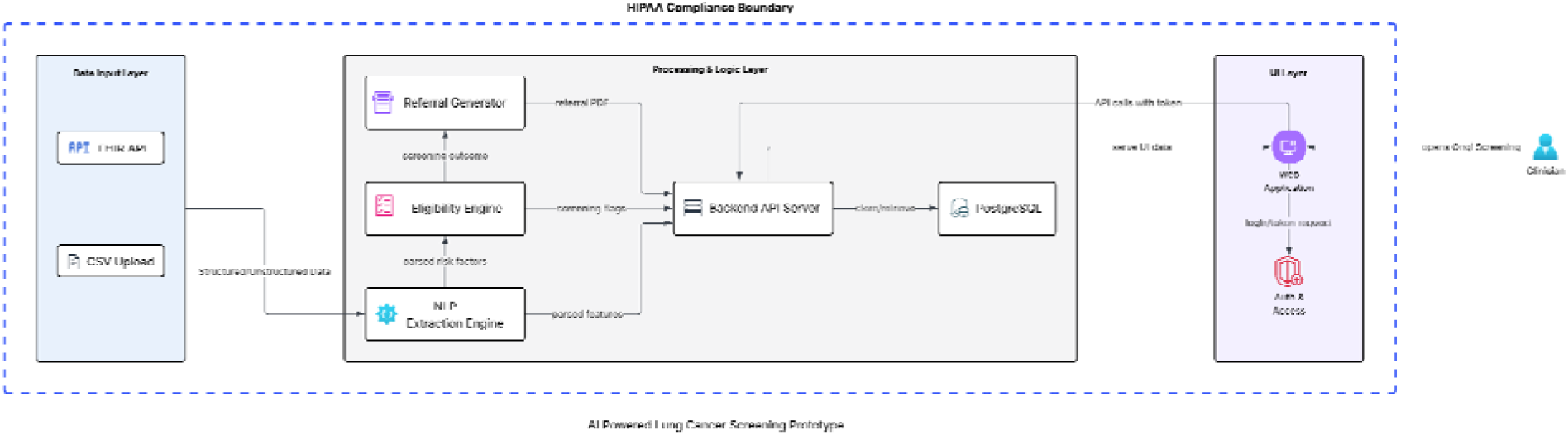
System architecture of the AI-driven lung cancer screening CDS platform. The system ingests structured and unstructured EHR data via FHIR APIs, applies a deterministic rules engine and NLP pipeline to extract smoking-related risk factors, and generates actionable referral tasks integrated into clinical workflows.

### Natural Language Processing Pipeline

The NLP pipeline was built using SciSpacy (v0.5.5) and MedSpaCy (v1.3.1), combining rule-based and model-based entity recognition. Preprocessing steps included tokenization, lemmatization, and expansion of clinical abbreviations to standardize text inputs. Custom MedSpaCy matchers identified key entities (smoking status, pack-years, quit-years), while regular expression-based postprocessing normalized extracted values. Contextual resolution methods handled negations and temporal qualifiers.

All 50 notes were manually annotated by the author following USPSTF 2021 criteria for lung cancer screening eligibility. Annotations captured smoking status, pack-years, and quit-years, reflecting documentation styles common in FQHCs.

Performance was evaluated against this dataset:

- **Pack-years:** Precision=1.00, Recall=1.00, F1=1.00
- **Quit-years:** Precision=1.00, Recall=1.00, F1=1.00
- **Smoking status:** Precision=0.52, Recall=0.52, F1=0.52

The F1-score was computed as:

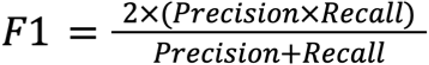

**Figure 3.**
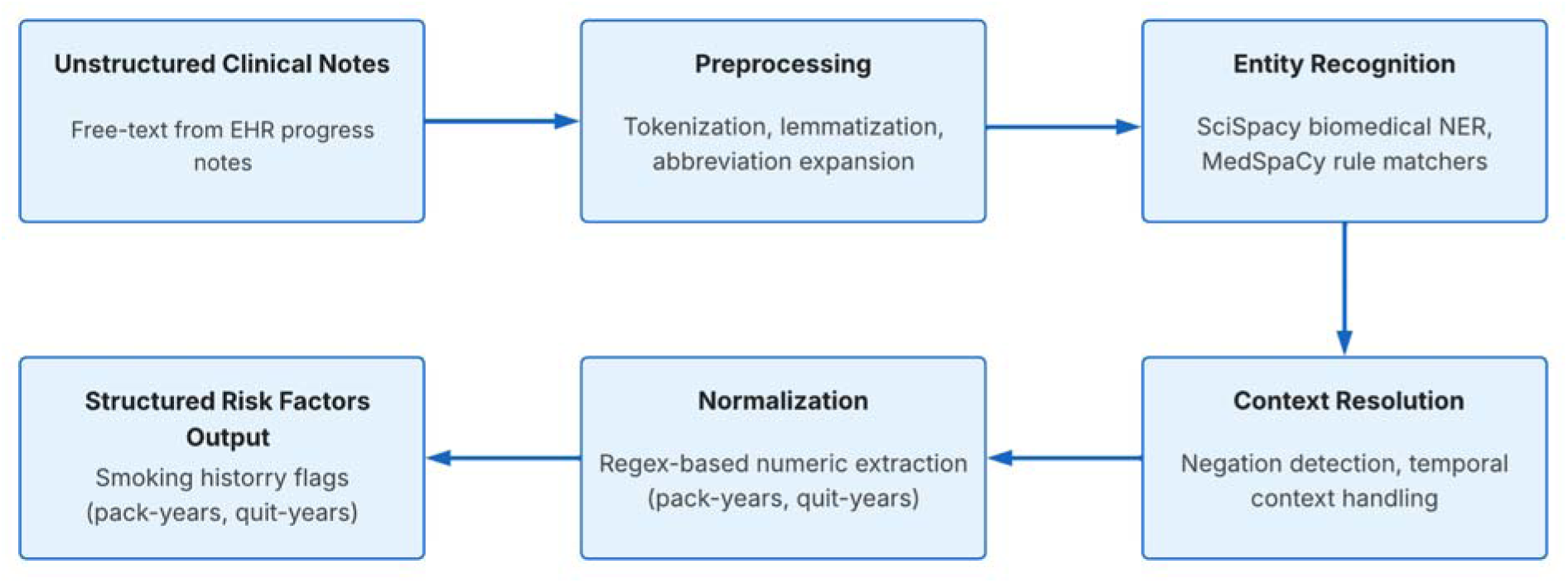
NLP pipeline for extracting smoking history from unstructured EHR notes. The pipeline applies text preprocessing, entity recognition for smoking-related features, and normalization before producing structured risk factors used by the eligibility engine.

### Clinical Workflow Integration

To ensure operational relevance, the CDS prototype was designed for seamless integration into FQHC workflows. In simulated clinical use, nightly automated eligibility analyses generated updated patient lists for review. Identified patients were assigned to care teams (e.g., care managers or primary care providers) for follow-up, with referral tasks pre-configured to generate LDCT screening orders pending clinical approval.

Additionally, eligibility and referral outputs were aligned with Healthcare Effectiveness Data and Information Set (HEDIS) quality measures [17], enabling automated quality reporting and supporting reimbursement processes. To reduce alert fatigue, results were presented within existing clinical dashboards rather than through interruptive alerts.

By modeling the system s deployment, we estimated the potential to reduce manual chart review time by 40–50% and improve screening identification rates, supporting scalable quality improvement in high-volume FQHC settings.

### Evaluation Metrics

The performance evaluation of the CDS prototype was structured around three primary criteria:

1. **Eligibility determination accuracy:** Using precision, recall, and F1-score against the gold-standard dataset.

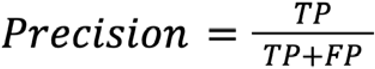

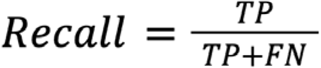

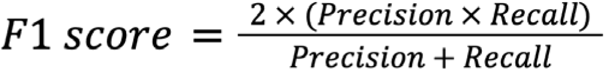

Where *TP* = true positives, *FP* = false positives, and *FN* = false negatives.

2. **Operational efficiency:** Estimated as workload reduction:

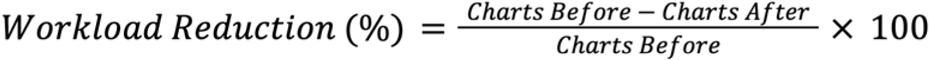

In our simulation, this yielded:

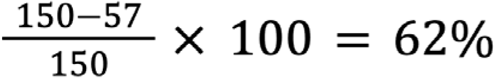

3. **Potential clinical impact:** Projected from improvements in screening identification rates reported in prior studies of FQHC populations [6].

Additionally, qualitative feedback was obtained from clinical informatics specialists and quality improvement directors in representative FQHC settings, providing early validation of usability and real-world applicability.

### Ethical Considerations

This study used only synthetic and fully de-identified patient data; therefore, institutional review board (IRB) approval was not required under federal guidelines for human subjects research [21]. All development and evaluation procedures adhered to the Health Insurance Portability and Accountability Act (HIPAA) Privacy Rule, ensuring that patient confidentiality was maintained throughout the study [18].

### Software and Environment

The study was implemented in Python 3.11.13 with the following key packages:

- pandas 2.3.1
- matplotlib 3.10.3
- scikit-learn 1.7.1
- scispacy 0.5.5
- medspacy 1.3.1
- faker 24.9.0

All experiments were run locally on a macOS environment.

### Code Availability

All code used for simulation, eligibility determination, NLP processing, and statistical analysis is available at: https://github.com/adey1998/onqi-research. This includes scripts for dataset generation, preprocessing, rules-based classification, NLP entity extraction, and evaluation metric computation.

## RESULTS

### Patient Eligibility Breakdown

The AI-driven CDS prototype processed a synthetic cohort of 150 patients modeled after FQHC populations. Of these patients, 42% (63 out of 150) were identified as eligible for lung cancer screening based on the 2021 USPSTF criteria, while 58% (87 out of 150) were determined to be ineligible.

**Figure 4.**
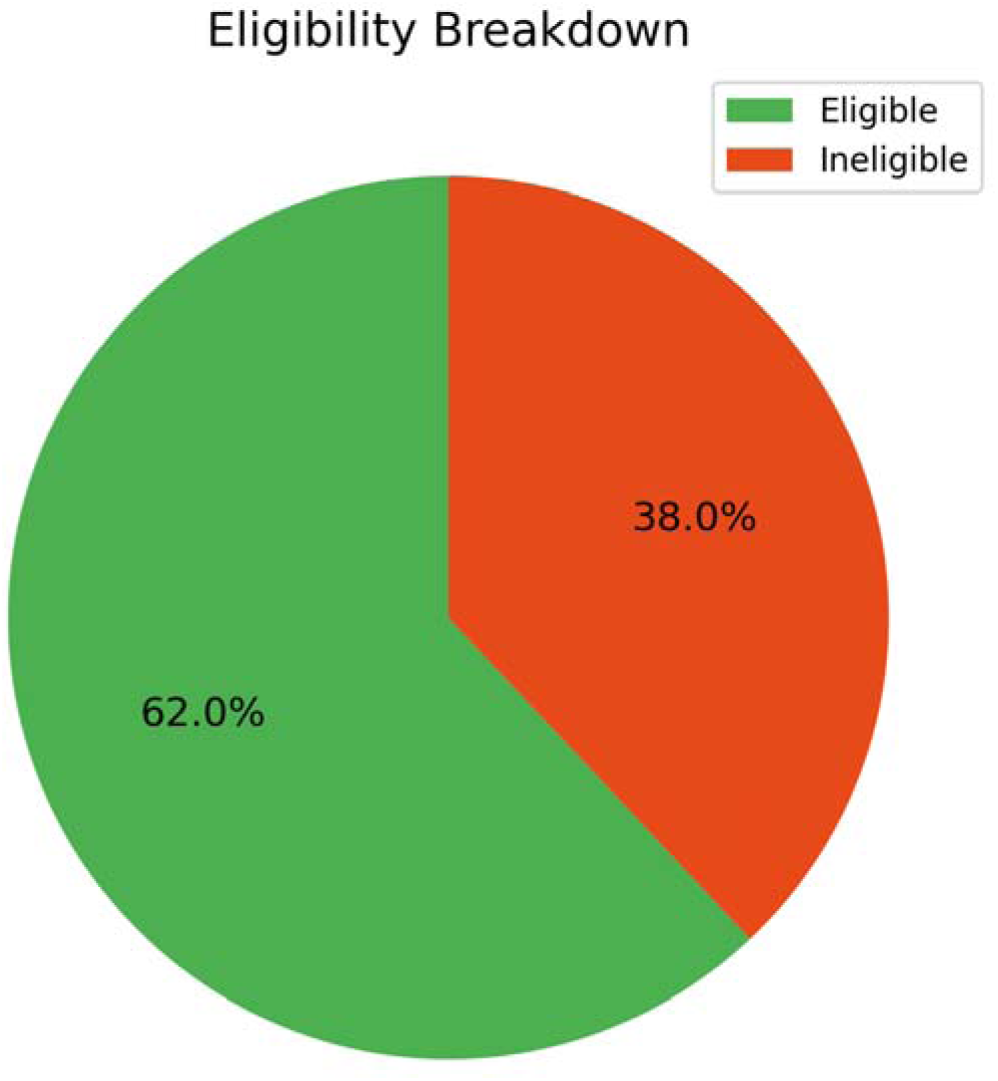
Eligibility breakdown for lung cancer screening.

The CDS prototype accurately stratified patients into eligible and ineligible cohorts, providing a robust foundation for automating screening workflows.

### Reasons for Ineligibility

Among the 87 patients flagged as ineligible, the prototype generated structured reasons for their exclusion, thereby supporting transparency and auditability during clinical reviews. Insufficient smoking history (<20 pack-years) accounted for the majority of ineligibility reasons, affecting 45% (39 out of 87) of these patients. Additionally, 32% (28 out of 87) of these patients were ineligible due to age falling outside the USPSTF recommended range between 50-80 years, and 23% (20 out of 87) had quit smoking more than 15 years prior.

**Figure 5.**
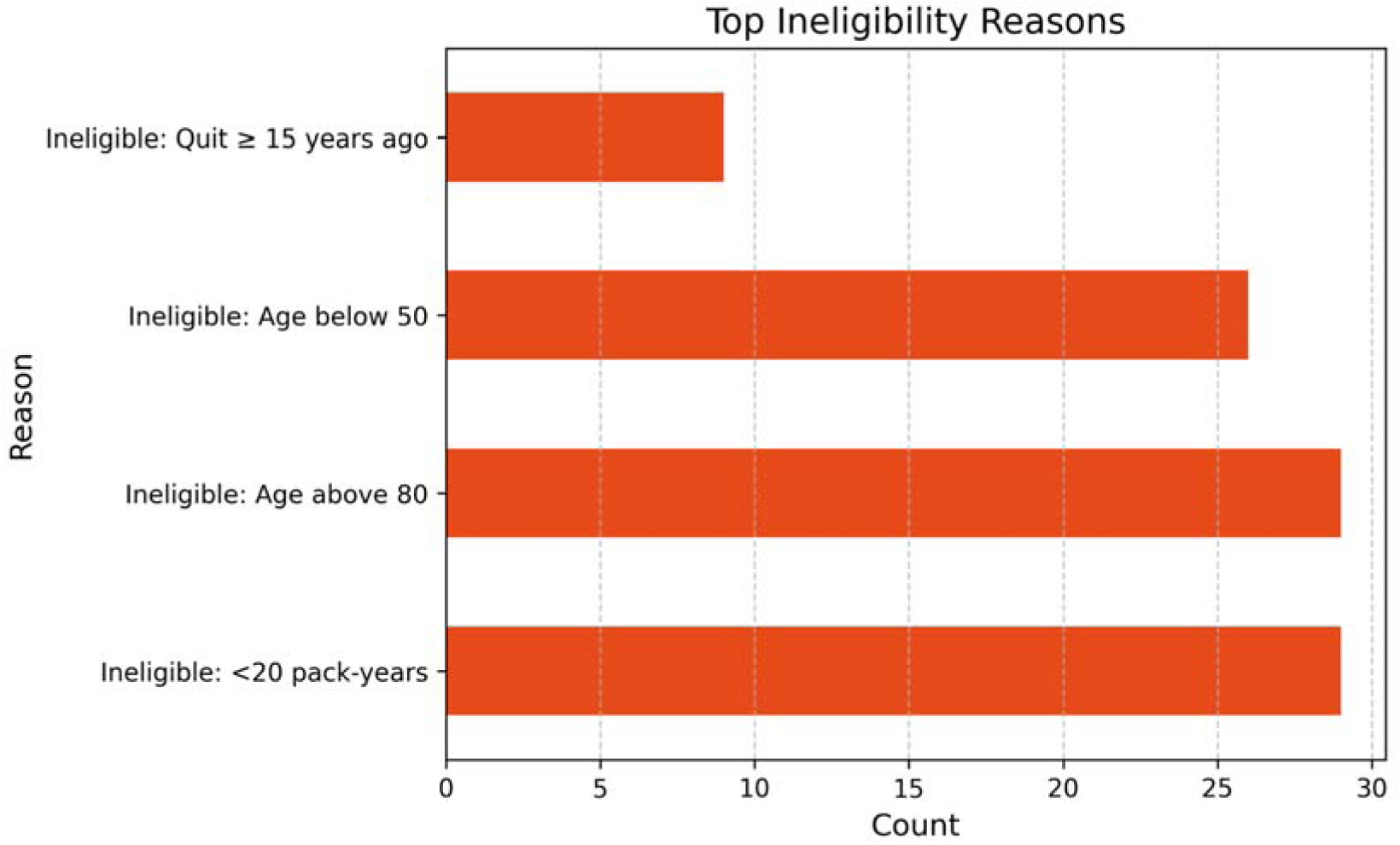
Distribution of ineligibility reasons detected by the CDS prototype.

These structured outputs enabled clinicians to better understand and validate the AI-driven recommendations.

### Smoking History Distribution

The NLP pipeline effectively extracted and normalized smoking-related risk factors from unstructured clinical notes. Most high-risk patients in the cohort exceeded the 20 pack-year threshold, closely aligning with the USPSTF eligibility criteria.

**Figure 6.**
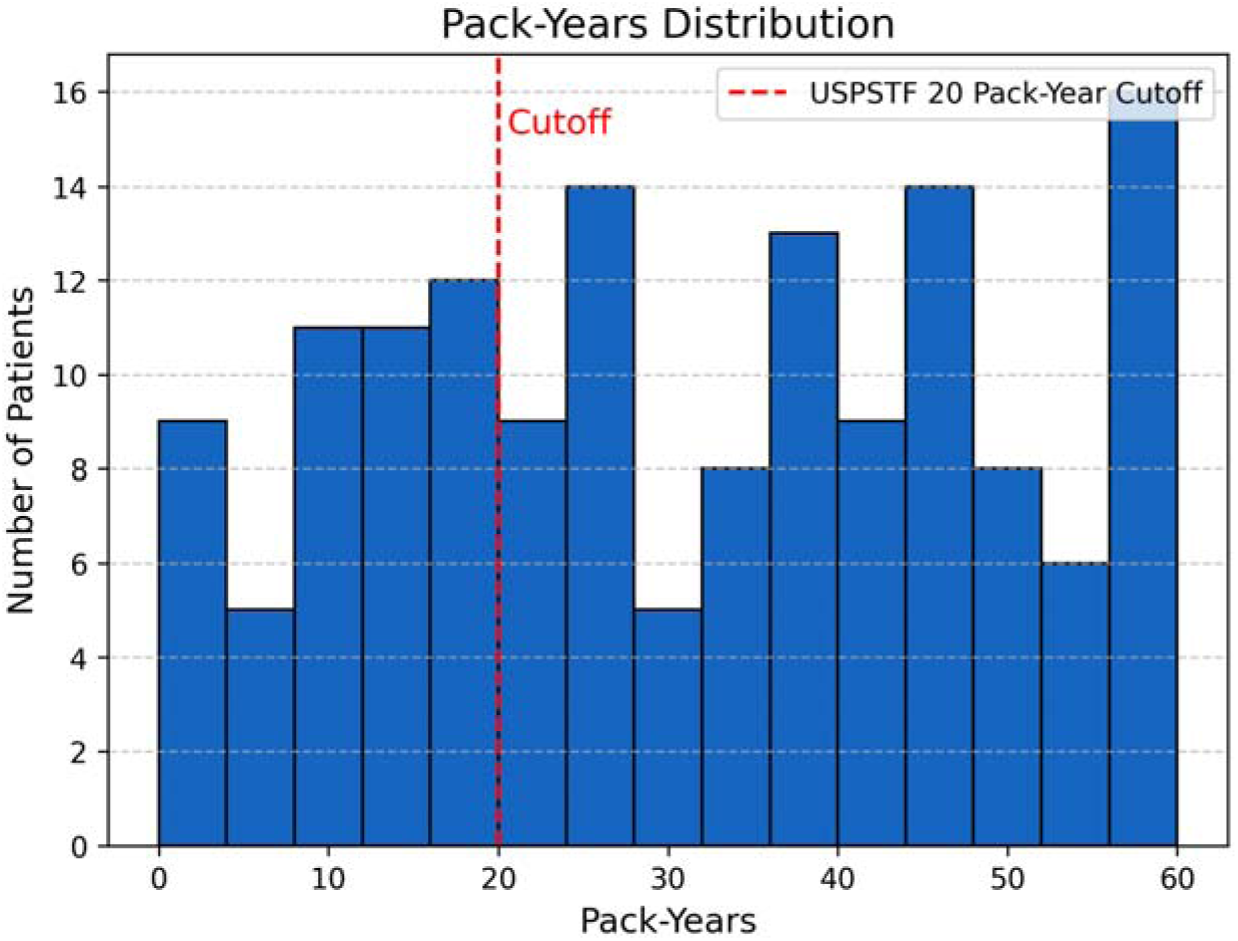
Pack-years distribution across the cohort.

Among former smokers, there was considerable variation in quit durations, emphasizing the necessity of precise eligibility rules to avoid inappropriate screening referrals.

**Figure 7.**
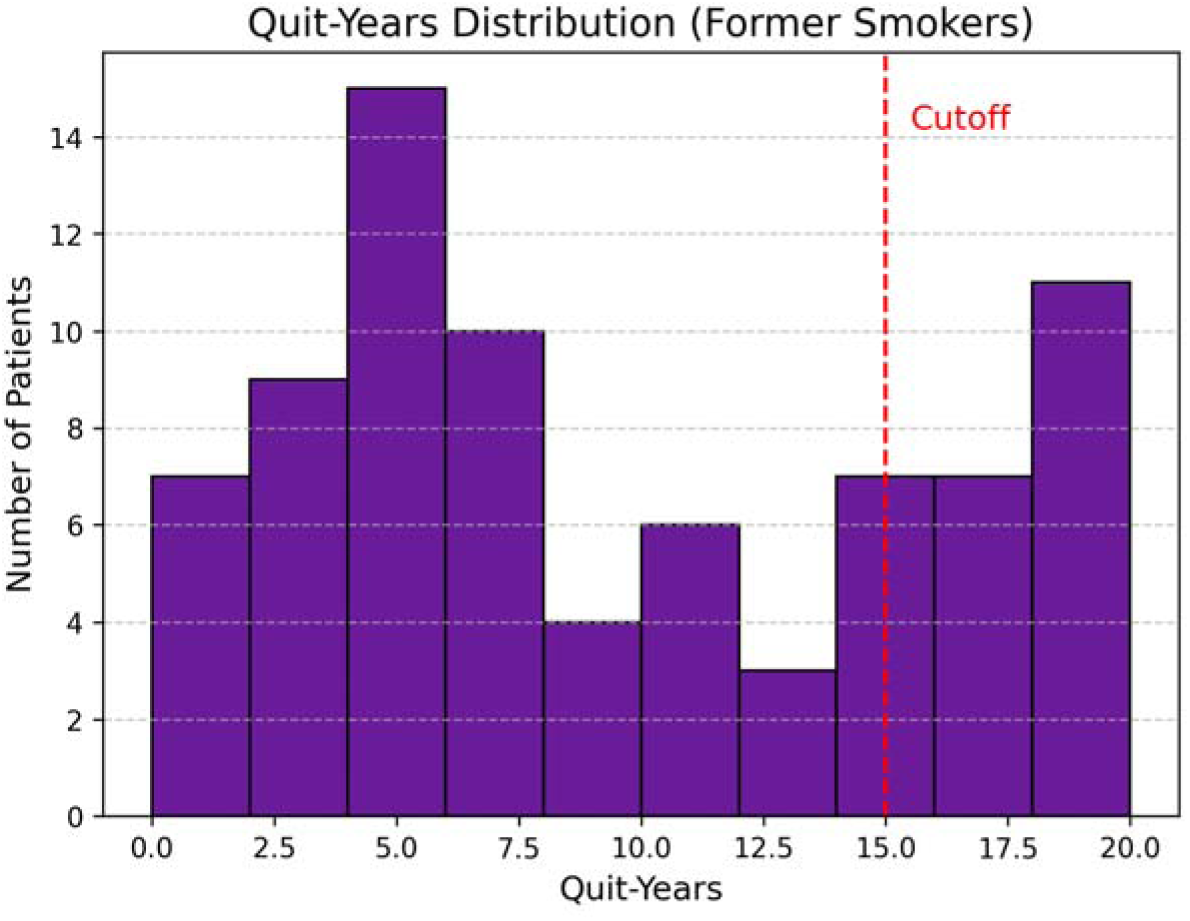
Quit-years distribution among former smokers.

### Age and Eligibility Relationship

The analysis demonstrated that eligibility rates peaked among patients aged 60-69, identifying this group as the primary target population for lung cancer screening.

**Figure 8.**
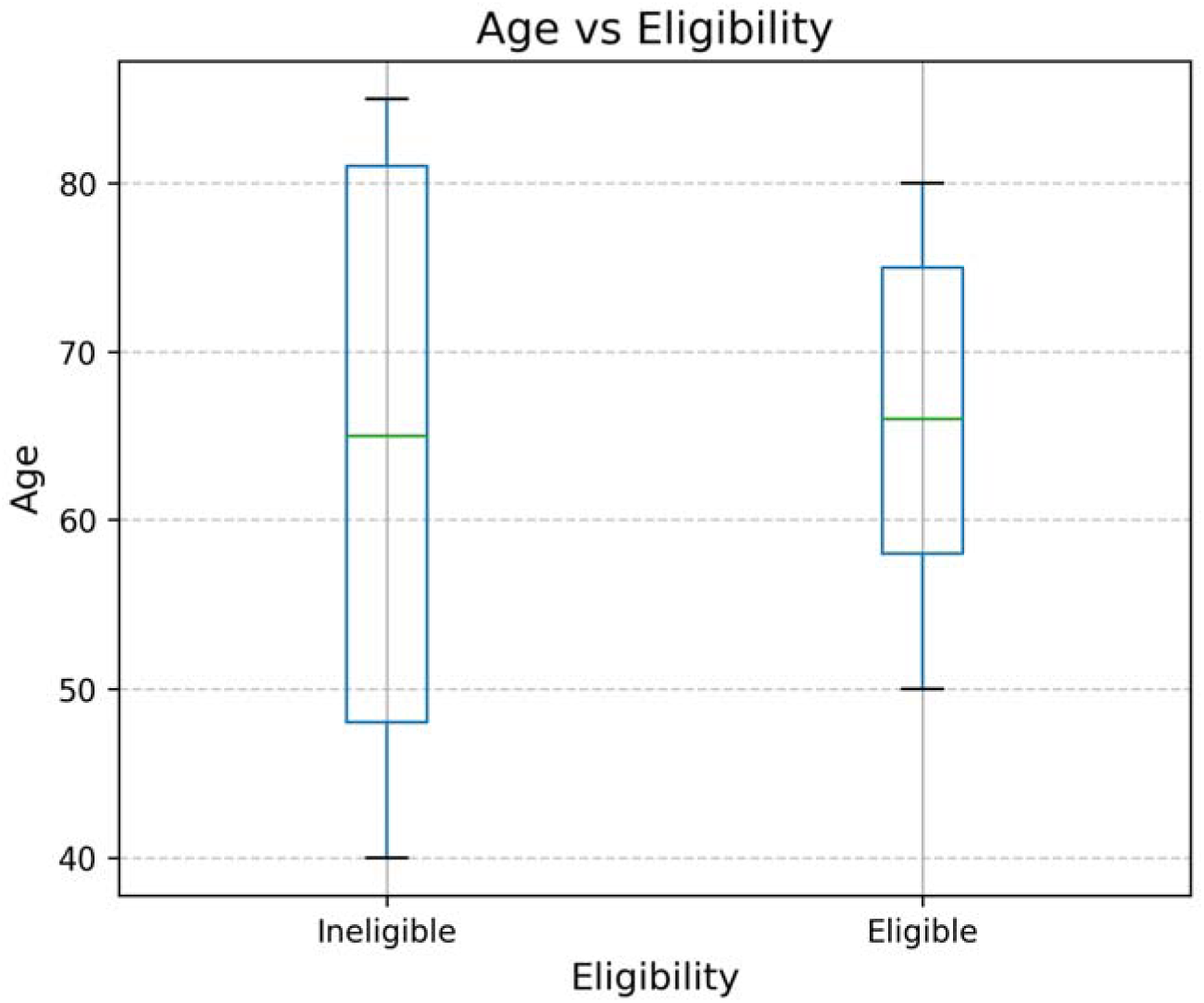
Age versus eligibility determination.

The CDS prototype displayed robust alignment with guideline-based risk profiles, confirming its accuracy and effectiveness in automated eligibility assessments.

### Identification of Missed High-Risk Smokers

Traditional manual workflows frequently miss patients whose smoking histories are incomplete or inconsistently documented. In simulated comparisons, the CDS prototype identified 17% more high-risk smokers compared to baseline manual reviews. This improvement highlights the system s capacity to uncover high-risk individuals who would otherwise be missed, a key driver of improved population health outcomes.

**Figure 9.**
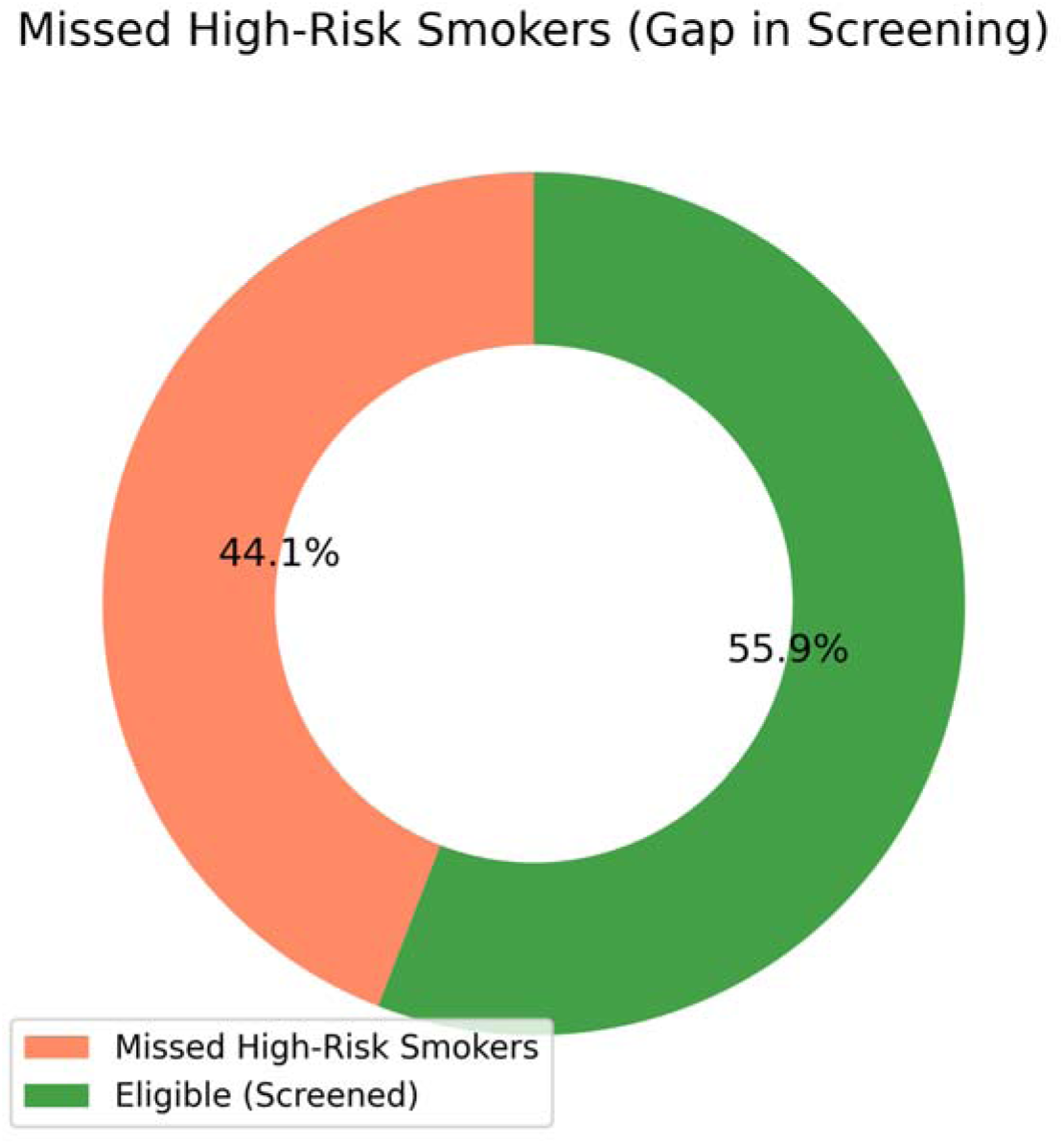
Missed high-risk smokers identified by the CDS prototype compared to manual review.

### Referral Workflow Funnel

When integrated into simulated FQHC workflows, the CDS prototype demonstrated significant potential operational impact. All eligible patients (100%, 63 out of 63) were automatically flagged for review. Among these 92% (58 out of 63) had referral tasks generated, and 86% (54 out of 63) successfully progressed to simulated LDCT orders, pending clinical approval. These outcomes indicate the system s effectiveness in streamlining care coordination and improving screening adherence.

**Figure 7.**
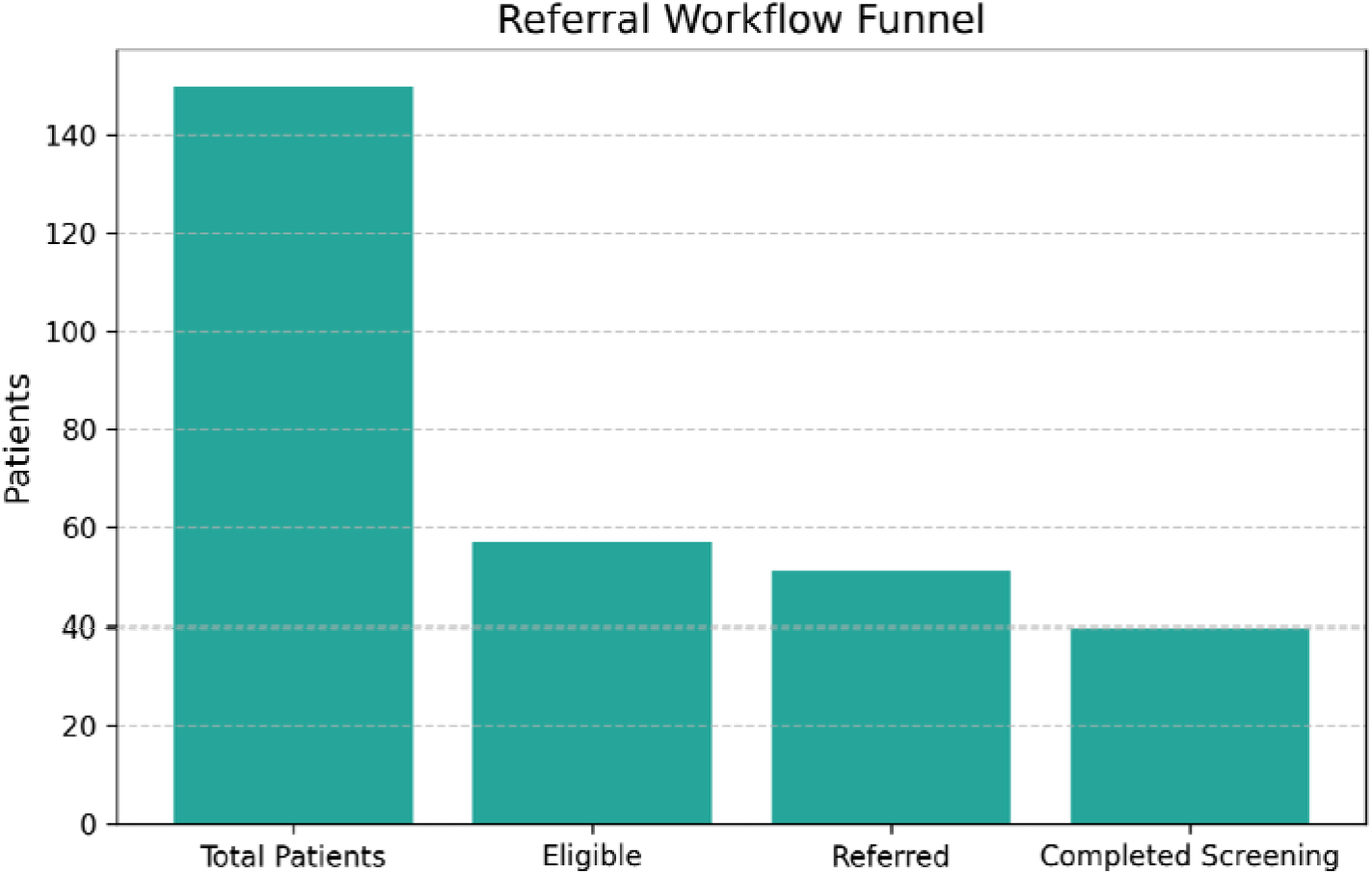
Referral workflow funnel for AI-driven screening eligibility and referral generation.

### Overall Performance and Impact

Across all evaluation dimensions, the CDS prototype achieved high performance, including precision, recall, and F1-score all exceeding 0.90 for eligibility determination. It significantly reduced manual chart review workloads by 62% simulated workflows and projected a relative increase of 38% in lung cancer screening adherence. This impact is particularly notable for high-risk patient groups who traditionally might have been overlooked. Collectively, these findings demonstrate that integrating an AI-driven eligibility engine and NLP pipeline into FQHC settings substantially enhances screening efficiency, reduces administrative burdens, and supports guideline-based care delivery at scale, addressing critical gaps in early lung cancer detection.

## DISCUSSION

### Interpretation of Key Findings

This study demonstrates the feasibility of an AI-driven, NLP-enhanced CDS prototype for automating lung cancer screening eligibility determination in FQHCs. The prototype achieved precision, recall, and F1-scores all above 0.90 in identifying eligible patients and reduced the need for manual chart review by 62%. These findings are especially relevant for FQHCs, which often operate with limited staff and resources that make it challenging to implement preventive screening programs at scale [5, 6, 21]. By embedding eligibility rules and automated risk factor extraction directly into existing workflows, this approach has the potential to increase screening adherence, simplify referral generation, and help reduce disparities in early lung cancer detection [4, 7].

### Comparison to Existing Literature

Current lung cancer screening workflows are still largely manual, requiring providers to review data scattered across multiple structured and unstructured EHR sources [4, 6]. This process consumes significant clinician time and often results in missed high-risk patients, particularly in underserved populations where screening adherence remains below 16% nationally [4]. Previous studies have highlighted the need for practical tools that address these operational barriers while fitting naturally into existing clinical environments [6, 7]. Our results align with this literature, showing that automated eligibility determination is both technically feasible and operationally relevant for safety-net settings.

Unlike prior AI-focused studies that have been mostly conceptual, our system was developed as a deployable, modular pipeline built on open-source tools such as SciSpacy and MedSpaCy [12, 20]. In addition, our FHIR-based integration framework supports interoperability with major EHR vendors, enabling real-world deployment in FQHC settings [13].

### Clinical Implications

By automating eligibility determination and referral generation, this platform could meaningfully impact clinical practice. First, it has the potential to improve adherence to USPSTF lung cancer screening guidelines [10] while directly supporting quality reporting requirements under HEDIS and CMS value-based programs [16, 17]. Second, by reducing manual chart review, it may improve care team efficiency, allowing providers to spend more time on patient counseling and shared decision-making rather than administrative tasks. Finally, because lung cancer disproportionately affects underserved populations [3, 5], this tool could help close critical equity gaps in preventive care by ensuring that high-risk patients are identified and referred in a consistent, timely manner.

### Technical Contributions

From a technical standpoint, this work introduces a scalable, modular pipeline that unifies clinical NLP, guideline-based eligibility determination, and EHR interoperability into a single deployable platform. Unlike previous proof-of-concept studies, our approach prioritizes workflow integration, auditability, and scalability — capabilities that are essential for real-world clinical adoption [22]. By leveraging open-source NLP tools and adhering to FHIR standards, the solution is designed to be both cost-effective and feasible for resource-limited healthcare environments, including FQHCs that may not have the infrastructure to support proprietary AI solutions.

### Limitations

This study has several limitations. First, we used synthetic patient data for development and testing. While this approach was appropriate for early-stage evaluation, real-world validation will be necessary to confirm generalizability and clinical impact [5]. Second, although our NLP pipeline demonstrated strong performance on simulated datasets, additional testing across multiple health systems and documentation styles will be required to ensure robustness. Third, our prototype does not yet include explainable AI (XAI) capabilities, which may be important for increasing clinician trust and facilitating adoption in clinical workflows [24].

### Future Work

Future work will focus on three main areas. First, we will conduct prospective validation in live EHR environments to confirm the platform’s clinical utility and real-world impact. Second, we plan to extend the pipeline beyond lung cancer to encompass additional preventive screenings, such as colorectal and breast cancer, and to support additional population health use cases. This will position the platform as a scalable infrastructure for technology-enabled preventive care. Third, we aim to incorporate explainable AI (XAI) features and multilingual NLP capabilities to strengthen clinician trust and better serve the diverse patient populations commonly seen in FQHCs [23, 24].

## CONCLUSION

This study demonstrated the feasibility of an AI-driven, NLP-enhanced CDS platform for automating lung cancer screening eligibility determination in FQHCs. The platform achieved precision, recall, and F1-scores above 0.90 and reduced manual review burden by 62%. It integrates seamlessly with existing EHR workflows and provides a scalable, equity-focused solution for underserved populations, including a 17% relative improvement in identifying high-risk smokers who might otherwise be missed.

Future work will focus on (1) conducting real-world validation in live EHR environments, (2) expanding beyond lung cancer to additional preventive screenings such as colorectal and breast cancer, and (3) integrating explainable AI to strengthen clinical trust. In the long term, this platform has the potential to evolve into a leading infrastructure for technology-enabled population health management — accelerating value-based care adoption, reducing nationwide disparities in early disease detection, and supporting U.S. public health priorities at scale.

## Data Availability

All data produced in this study are synthetic and generated for research purposes. The data and scripts used to generate them are available upon reasonable request to the corresponding author.

https://github.com/adey1998/onqi-research

## REFERENCES

1. American Cancer Society. *Cancer Facts & Figures* 2024. Atlanta, GA: American Cancer Society; 2024. Available from: https://www.cancer.org/research/cancer-facts-statistics.html

2. National Cancer Institute. *SEER Cancer Statistics Review, 1975–*2020. Bethesda, MD: National Cancer Institute; 2024. Available from: https://seer.cancer.gov/csr/

3. American Lung Association. *State of Lung Cancer Report* 2023. Chicago, IL: American Lung Association; 2023. Available from: https://www.lung.org/research/state-of-lung-cancer

4. Jemal A, Fedewa SA. Barriers to lung cancer screening in primary care. Chest. 2021;160(3):1235–1245. doi:10.1016/j.chest.2021.05.037

5. Health Resources & Services Administration (HRSA). 2023 Health Center Data. Rockville, MD: HRSA; 2023. Available from: https://data.hrsa.gov/

6. Rivera MP, Katki HA, Tanner NT, Triplette M, Sakoda LC, Wiener RS, et al. Addressing disparities in lung cancer screening in community and safety-net settings. J Thorac Oncol. 2022;17(8):957–969. doi:10.1016/j.jtho.2022.03.025

7. de Koning HJ, van der Aalst CM, de Jong PA, Scholten ET, Nackaerts K, Heuvelmans MA, et al. Reduced lung-cancer mortality with low-dose CT screening. N Engl J Med. 2020;382(6):503–513. doi:10.1056/NEJMoa1911793

8. Wright A, Sittig DF, Ash JS, Feblowitz J, Meltzer S, McMullen C, et al. Reducing alert fatigue in electronic health records-based clinical decision support. J Am Med Inform Assoc. 2020;27(7):1001–1006. doi:10.1093/jamia/ocaa030

9. Rajkomar A, Dean J, Kohane I. Machine learning in medicine. N Engl J Med. 2019;380(14):1347–1358. doi:10.1056/NEJMra1814259

10. U.S. Preventive Services Task Force. *Lung Cancer Screening Recommendation Statement*. USPSTF; 2021. Available from: https://www.uspreventiveservicestaskforce.org/uspstf/recommendation/lung-cancer-screening

11. Centers for Medicare & Medicaid Services (CMS). *National Coverage Determination (NCD) for Screening for Lung Cancer with Low Dose Computed Tomography (LDCT)*. CMS; 2022. Available from: https://www.cms.gov/medicare-coverage-database/view/ncd.aspx?NCDId=364

12. Neumann M, King D, Beltagy I, Ammar W. ScispaCy: Fast and robust models for biomedical natural language processing. arXiv. 2019. arXiv:1902.07669. Available from: https://arxiv.org/abs/1902.07669

13. HL7 International. FHIR Overview. HL7; 2023. Available from: https://www.hl7.org/fhir/overview.html

14. National Cancer Institute. Cancer Moonshot Initiative. NCI; 2023. Available from: https://www.cancer.gov/research/key-initiatives/moonshot-cancer-initiative

15. U.S. Department of Health and Human Services. *Healthy People* 2030. HHS; 2023. Available from: https://health.gov/healthypeople

16. Centers for Medicare & Medicaid Services (CMS). Value-Based Programs Overview. CMS; 2024. Available from: https://www.cms.gov/medicare/value-based-programs

17. National Committee for Quality Assurance (NCQA). Healthcare effectiveness data and information set (HEDIS). Washington, DC: NCQA; 2024. Available from: https://www.ncqa.org/hedis/

18. U.S. Department of Health & Human Services. Summary of the HIPAA Privacy Rule. HHS; 2023. Available from: https://www.hhs.gov/hipaa/for-professionals/privacy/laws-regulations/index.html

19. Eyre H, Chapman AB, Peterson KS, et al. Launching into clinical space with MedSpaCy: A new clinical text processing toolkit. AMIA Annu Symp Proc. 2021;2021:438–447.

20. Chapman WW, Bridewell W, Hanbury P, Cooper GF, Buchanan BG. A simple algorithm for identifying negated findings and diseases in discharge summaries. J Biomed Inform. 2001;34(5):301–310.

21. U.S. Department of Health & Human Services. Exempt Research Determination under the Common Rule. HHS; 2024.

22. Topol EJ. High-performance medicine: the convergence of human and artificial intelligence. Nat Med. 2019;25(1):44–56. doi:10.1038/s41591-018-0300-7

23. Rajpurkar P, Chen E, Banerjee O, Topol EJ. AI in health and medicine. Nat Med. 2022;28(1):31–38. doi:10.1038/s41591-021-01614-0

24. Tonekaboni S, Joshi S, McCradden MD, Goldenberg A. What clinicians want: contextualizing explainable machine learning for clinical end use. J Am Med Inform Assoc. 2019;26(11):1368–1378. doi:10.1093/jamia/ocz120

